# Talking while Walking After Concussion: Acute effects of concussion on speech pauses and gait speed

**DOI:** 10.1101/2024.08.06.24311478

**Authors:** Shu Yang, Paula K. Johnson, Colby R. Hansen, Elisabeth A. Wilde, Melissa M. Cortez, Leland E. Dibble, Peter C. Fino, Tiphanie E. Raffegeau

**Author notes:** Corresponding Author: Peter C. Fino, PhD, 250 S 1850 E, Rm 257, Salt Lake City, UT, 84112, USA. Denotes Co-Senior authorship (Peter C. Fino, Tiphanie E. Raffegeau).

## Abstract

**Background:** Deficits in dual-tasks (DT) are frequently observed post-concussion (i.e., mild Traumatic Brain Injury). However, traditional DT may not be relevant to daily life. Walking while talking elicits DT costs in healthy adults and is part of daily life.

**Objective:** We investigated the effect of concussion on walking with extemporaneous speech and explored relationships between DT and acute symptoms.

**Methods:** Participants with recent concussion (<14 days post-injury) and controls completed three tasks: single-task gait without speaking (ST_G_), single-task speaking without walking (ST_S_) and walking while speaking (DT). Silent pauses in speech audio reflected cognitive performance, and gait was quantified using inertial sensors. We used linear mixed models to compare groups and conditions and explored associations with self-reported symptoms.

**Results:** Both concussion (n=19) and control (n=18) groups exhibited longer speech pauses (*p* < 0.001), slower walking speeds (*p* < 0.001), and slower cadence (*p* < 0.001) during the DT compared to ST conditions. There were no group differences or interactions for speech pauses (*p* > 0.424). The concussion group walked slower (*p* = 0.010) and slowed down more during DT than the control group (group*task *p* = 0.032). Vestibular symptoms strongly associated with ST speech pause duration (*ρ* = 0.72), ST gait speed (*ρ* = −0.75), and DT gait speed (*ρ* = −0.78).

**Conclusions:** Extemporaneous speech is well-practiced, but challenging to complete while walking post-concussion. Strong associations between DT outcomes and vestibular-related symptoms suggest DT deficits vary with post-concussion symptomology. DT deficits may be deleterious to daily tasks post-concussion.

## INTRODUCTION

Cognitive and motor problems, such as balance and gait deficits, are common following concussions ^1–3^. Performing cognitive-motor dual-tasks (DT) – completing simultaneous cognitive and motor tasks at the same time – elicits slower gait speeds, shorter strides, and longer stride times in people after a concussion ^2,4–7^. DT deficits arise from an inability to handle competing cognitive and motor demands and are thought to result from deficits in attentional control and a loss of automaticity – defined as an increased reliance on cognitive resources for relatively automatic tasks (e.g., walking) ^8,9^. After a concussion, DT deficits can remain for months and persist long after the resolution of other signs and symptoms ^5,10,11^. Further, DT deficits are reported in people with chronic, persisting post-concussion symptoms; people with persisting concussion-related symptoms prioritize the cognitive task, exhibiting higher costs to gait performance than cognitive task performance while carrying out both simultaneously, referred to as a “posture second” strategy ^12,13^. The ‘posture second’ is a compensation that reflects an inability to appropriately allocate resources to motor performance when faced with a cognitive challenge ^14,15^. Yet, the consequences of DT deficits during daily life remain unclear because prior work has typically studied cognitive tasks that are irrelevant to daily life ^16^.

Ideally, studies of cognitive-motor DT effects should quantify both the cognitive and the motor task with equivalent resolution to examinate the potential for bidirectional interference ^17^. Unfortunately, prior DT paradigms have used contrived arithmetic, alphabet recitation, memorization, visuospatial, and auditory/visual Stroop tasks when testing concussion populations that are not measured continuously ^16^. Although these tasks are easy to administer and score, they may not represent the cognitive demands of everyday life, like walking and talking. Further, laboratory-based cognitive tests may be affected by individual differences in education/socio-economic background, mathematics-related anxiety or skill-level, comfort with public speaking, and task-engagement ^18^. Typical contrived tasks are vulnerable to compensatory strategies that may not reflect the function of targeted cognitive behaviors ^19^, and boast learning and practice effects that are difficult to control for in an experimental setting ^20^.

In contrast to traditional arithmetic or recitation tasks, extemporaneous speech is a cognitive task that is highly practiced, relevant to daily living, commonly performed during other motor tasks such as walking, and elicits DT costs in healthy young adults ^18,21–23^. Extemporaneous speech involves complex cognitive processes (e.g. language formulation, executive function, processing speed) and demands ongoing cognitive processing and continuous language retrieval. Similar to gait, the cognitive load of extemporaneous speech can be obtained from measures of speed and fluency; speech rate per syllable decreases under high cognitive-linguistic demands, with longer and more prevalent silent pauses ^24^. A natural pause during extemporaneous speech is around 150 ms to 250 ms ^25^, and longer silent speech pauses serve as an indicator of language fluency ^26,27^. Silent pauses are associated with cognitive planning and memory retrieval and reflect syntactic complexity in language formulation, and more frequent pauses at prosodic boundaries (i.e. clauses, sentence breaks) are associated with reduced syntactic complexity ^27^. Therefore, the frequency and duration of silent pauses during extemporaneous speech reflect cognitive-linguistic demands ^27,28^.

Speech deficits such as slower articulation rate have been documented in moderate-to-severe traumatic brain injuries (TBI) ^29^ and speech pauses may be an indicator of mild TBI (i.e., concussion). One study observed individuals exhibited more pauses and filler words during a picture description task acutely after concussion compared to their pre-concussion performance ^30^. Separately, a machine learning-based analysis of speech patterns revealed high diagnostic accuracy in a small preliminary sample of athletes after concussion ^31^. Further, clinical care patterns highlight the role of speech and language deficits post-concussion; a chart review indicated 43% of pediatric patients were referred from a specialty concussion clinic to speech language pathologists to treat issues with communication, attention, and memory ^32^. Yet, it remains unclear how these concussion-related deficits in speech fluency, defined here as the frequency and duration of silent pauses in extemporaneous speech, interact with competing motor tasks, such as walking.

The purpose of our study was to investigate the effect of concussion on extemporaneous speech production during single-task (ST) and DT walking paradigms. Our primary hypotheses were that people recovering from concussion would exhibit impaired speech production, defined by more frequent and longer silent pauses during speech; that both groups would exhibit a decline in speech production when walking compared to sitting; and that those with a concussion would exhibit greater DT costs (DTC) to gait and speech compared to the control group, defined as slower DT gait speed or longer and more frequent DT speech pauses. As a secondary aim, we explored the association between self-reported post-concussion symptoms and the DT effects of walking and talking. As extemporaneous speech production is understudied in people with concussion, we also explored the associations between traditional neurocognitive tests with ST and DT speech and gait performance outcomes using the National Institutes of Health (NIH) Cognition Toolbox.

## MATERIALS and METHODS

### Participants

As part of a larger study, 22 participants with recent concussions (<14 days post-injury) and 19 healthy control participants were recruited and provided informed written consent for the study. All protocols were approved by the Institutional Review Board, and participants provided informed written consent in advance. Concussion participants were identified and recruited using electronic medical records indicating a recent concussion-related injury. Control participants were recruited from the local community using flyers, online postings, and public dissemination, and were age- and gender-matched to the concussion participants. Candidates were included if they had no history of neurological illness (e.g. stroke), history of a major neurological condition (e.g. epilepsy), major psychiatric disorders that required in-patient hospitalization, orthopedic conditions that would explain balance or gait issues, history of vestibular or orthostatic blood pressure problems, and no more than three concussions in their lifetime. Additionally, healthy control participants had no history of concussion or concussion symptoms in the five years prior to participation. Due to other components of the larger study, participants also had no contraindications for an MRI.

### Procedures

Participants completed a series of cognitive, balance, mobility, autonomic, and neuroimaging assessments as part of the larger study; however, the current analysis is focused on a subset of balance and cognitive tasks. First, participants completed the Neurobehavioral Symptom Inventory (NSI) to identify self-reported symptoms ^33^. Participants also completed the NIH-Toolbox Cognition battery as a computerized assessment of several aspects of cognition ^34^ including: Picture Vocabulary, Flanker Inhibitory Control and Attention, List Sorting, Dimensional Change Card Sort, Pattern Comparison, Picture Sequence Memory, Oral Reading Recognition. All NIH Toolbox tests were administered via an iPad with the test administrator in an isolated room. Three summary scores included the Total Composite, the Fluid Composite, which included the Dimensional Change Card Sort, Flanker Inhibitory Control and Attention, Picture Sequence Memory, List Sorting, and Pattern Comparison, and the Crystallized Composite, which included the Picture Vocabulary and Oral Reading Recognition tests.

The cognitive-motor assessment included three tasks: single-task speaking while seated (ST_S_), single-task gait without speaking (ST_G_), and walking while speaking (DT). By design, continuous measures of both cognitive and motor tasks with equivalent resolution were selected to gain insight into dual-task effects on both cognitive and motor function. Participants were given a list of pre-designed topics and asked to select five topics from a list of 20 that they felt most comfortable discussing (e.g., favorite childhood memory). Participants were instructed that they should select topics that they could talk about for one minute continuously and that it did not matter what they said, just that they kept talking the entire time. Next, participants were seated and given one prompt from the five selected topics (e.g., “Tell me about your favorite childhood memory”). A timer was used to ensure all participants spoke for one minute. Following the seated speech task (ST_S_), participants were instructed to walk back and forth between two lines spaced 20 meters apart at their comfortable pace for one minute (ST_G_). Following both ST conditions, participants completed a DT condition where they were assigned a new topic and instructed to repeat the walking task while talking about their given topic. Participants were not provided explicit instructions to prioritize either task.

Audio from each task was recorded using a lapel microphone and wireless transmitter (WMX-1, Movo Photo, Los Angeles CA, USA) connected to an iPad (8^th^ generation, Apple Inc.) such that audio and video were recorded simultaneously. Standard spatiotemporal measures of gait speed and cadence were obtained for each walking trial using inertial sensors (APDM Inc., Portland OR, USA) placed bilaterally on the feet, lumbar spine, forehead, and sternum. Inertial sensors recorded tri-axial acceleration and angular velocity, which was processed using validated, automated algorithms (Mobility Lab v2, APDM Inc. Portland OR, USA) to obtain gait speed and cadence ^35^. To account for differences in stature, gait speed was normalized by dividing the speed by participant height.

### Data Analysis

All audio recordings were trimmed to exactly one minute long, removing all noise before and after the cues to start and stop talking. Audio files were then imported into MATLAB and processed using a custom script to identify speech and measure silent pause frequency and duration for each participant. The audio signal for each recording was filtered using a 4^th^ order bandpass filter with a passband of 100 Hz to 5000 Hz to isolate frequencies and their harmonics associated with speech. A moving variance window of 50 ms was then applied to the signal, and a ‘speech threshold’ equal to the 40^th^ percentile of the windowed variance was used to identify the presence or absence of speech, based on agreement with manual identification of silent pauses. Durations longer than 250 ms without speech (windowed variance > 40^th^ percentile) were designated as silent speech pauses. The 250 ms threshold was selected based on prior work that indicated an increase in speech pauses were related to increased cognitive demand during walking ^29,30,36–38^. The total duration of silent pauses was selected as the primary outcome. Secondary outcomes included the total number of pauses.

### Statistical Analysis

To investigate the effect of concussion on extemporaneous speech production during ST and DT walking, we implemented linear mixed effect regression models for each speech and gait outcome. Models included fixed effects for group, task (single vs. dual), and the group*task interaction. Models were adjusted for covariates of age and sex. Random intercepts by subject were included to account for within-subject correlations across tasks. Between-group effect sizes were calculated for both single- and DT conditions using Hedges’s *g* ^39^. A significance level of 0.05 was used for each analysis.

To explore the associations between self-reported symptoms and neurocognition on the DT effects of walking and talking, Pearson correlation coefficients were computed between the primary outcomes (total pause duration and gait speed) and the symptom categories (affective, cognitive, somatosensory, vestibular, and total) of the Neurobehavioral Symptom Inventory (NSI) within the concussion group and fully adjusted t-scores from the NIH Toolbox Cognition battery from groups separately. In addition to primary outcomes during ST and DT conditions, DT costs (DTC) were calculated for both speech (DTC_S_) and gait (DTC_G_) outcomes using 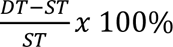, where ST and DT represent the ST and DT outcome. To ensure all outcomes remained in the same direction (i.e., more negative value indicates worse performance), all speech outcomes were multiplied by a negative sign for DTC_S_ calculations. Correlations were computed between self-reported symptoms and all ST, DT, and DTC outcomes. As this was an exploratory analysis, correlation coefficients were interpreted using guidelines proposed by Rowntree (Rowntree, 1981): values less than 0.2 indicate a very weak relationship, 0.20 to .39 indicate a weak relationship, 0.40 to 0.69 indicates a moderate relationship, 0.70 to 0.89 indicates a strong relationship, and 0.90 or greater indicates a very strong relationship. All analyses were conducted in MATLAB using the Statistics and Machine Learning Toolbox (r2020a, The MathWorks, Inc.).

## RESULTS

Speech recordings from four participants were excluded due to poor audio quality that prohibited analysis, leaving a total of 19 adults with concussion and 18 healthy controls included. Full demographic information is provided in Table 1.

**Table 1.** Demographic information about subjects in each group. All quantities are reported as mean (SD) unless otherwise noted.

|  |  |  |
| --- | --- | --- |
|  | Concussion | Control |
| N (M/F)* | 19 (15/4) | 18 (9/9) |
| Age (yrs) | 31.4 (9.3) | 27.7 (7.1) |
| Height (cm) | 178.1 (8.7) | 174.1 (12.8) |
| Mass (kg) | 80.5 (19.7) | 70.4 (12.4) |
| Race* |  |  |
| American Indian or Alaska Native | 0 | 0 |
| Asian | 1 | 3 |
| Black or African American | 0 | 0 |
| Native Hawaiian or Other Pacific Islander | 1 | 0 |
| White | 17 | 14 |
| More than One Race | 0 | 1 |
| Time since concussion (days) | 8.6 (3.1) | N/A |
| NSI Total Symptom Score | 31.6 (21.6) | 5.8 (6.0) |
| NSI-Affective | 9.2 (7.8) | 3.2 (3.9) |
| NSI-Vestibular | 4.6 (3.3) | 0.1 (0.3) |
| NSI-Cognitive | 7.4 (4.4) | 0.8 (1.2) |
| NSI-Somatosensory | 9.2 (6.3) | 1.4 (1.3) |
| * reported as count |  |  |
| + reported as median (range) |  |  |
| NSI = Neurobehavioral Symptom Inventory |  |  |

Both groups exhibited bidirectional interference based on the DTC_G_ and DTC_S_, though there was more variability in the DTC_S_ outcomes across participants (Figure 1). Both concussion and control groups exhibited longer total pause durations (*p* < 0.001), slower speeds (*p* < 0.001), and slower cadence (*p* < 0.001) during the DT compared to ST_G_ and ST_S_ conditions (Table 2). There was no significant difference in total speech pause duration between groups (*p* = 0.424) or significant group*task interaction (*p* = 0.941). However, there were notable between-group effect sizes for both ST_S_ and DT conditions (*g* = 0.42, *g* = 0.57, respectively; Table 2). The concussion group walked at slower speeds (*p* = 0.010) and there was a significant group*task interaction for gait speed (*p* = 0.032), indicating that participants with concussion walked slower than controls during both ST_G_ and DT, and they exhibited greater reductions in speed from ST_G_ to DT compared to control subjects. There was no group difference (*p* = 0.133) nor group*task interaction (*p* > 0.091) for cadence. No group, task, or group*task effects was detected for the number of pauses per minute, but notable between-group effect sizes were observed for the number of pauses in the DT condition (*g* = 0.57, Table 2).

**Figure 1.**
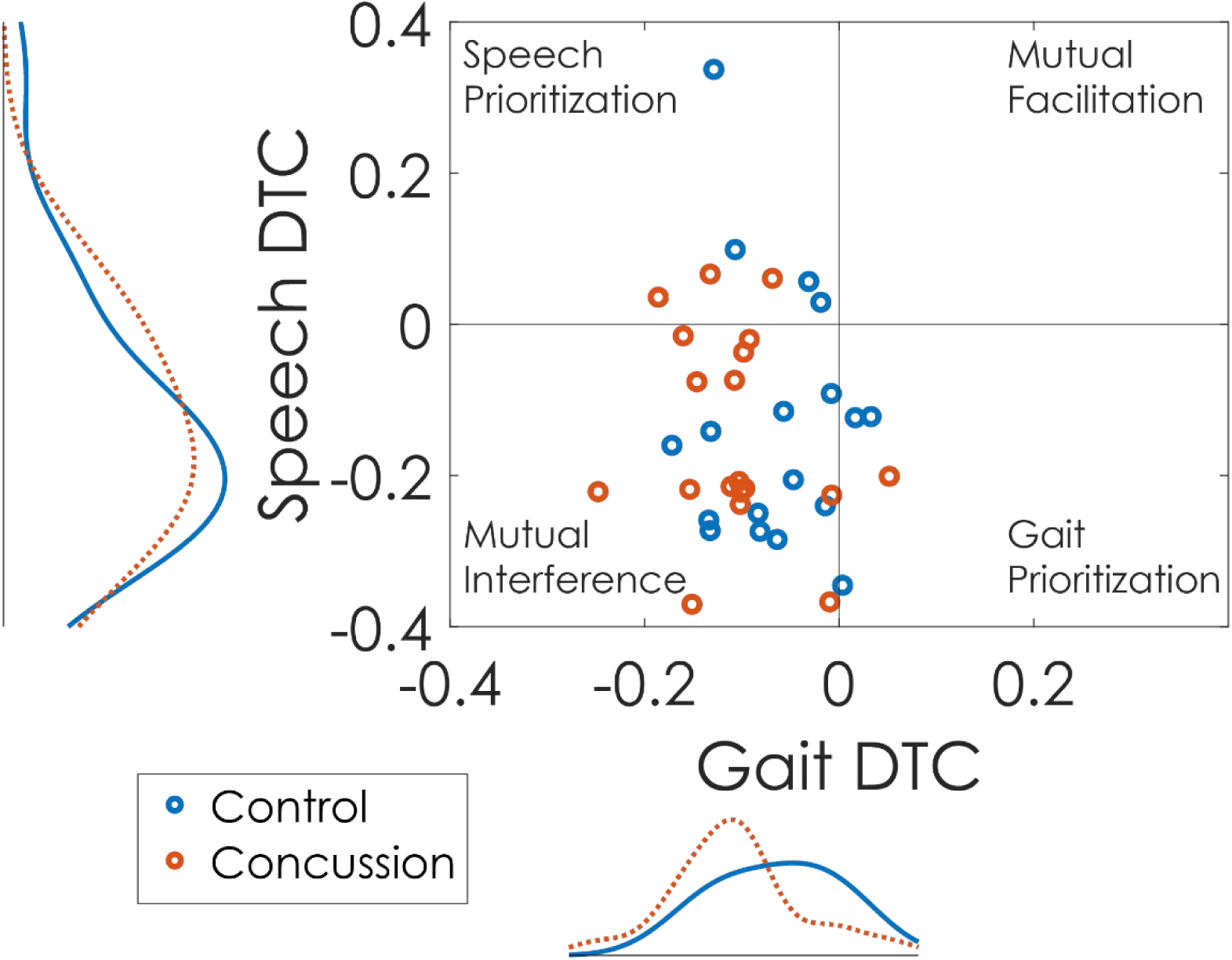
Scatter histograms depicting the dual-task (DT) effect for speech (y-axis) and gait (x-axis) as a percentage of single-task (ST) performance for participants with concussion (orange) and healthy controls (blue). Dual-task effects were calculated as 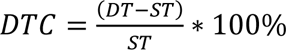. Kernal density plots are presented adjacent to the scatter plot indicating the distributions of each group.

**Table 2.** Descriptive statistics of speech and gait outcomes by group and task (single-task, ST and dual-task, DT). P-values are from the linear mixed models and represented contrast-coded effects when adjusting for sex and age. Gait speed was normalized to the participant height.

|  | Concussion |  | Control |  | P-value<br>(Group) | P-value<br>(Task) | P-value<br>(Group*Task) | Between-Group<br>Effect sizes<br>Hedges' *g* |  |
| --- | --- | --- | --- | --- | --- | --- | --- | --- | --- |
|  | ST | DT | ST | DT |  |  |  | ST | DT |
| **Speech Fluency** |  |  |  |  |  |  |  |  |  |
| Total Pause<br>Duration (s) | 14.2<br>(2.6) | 16.1<br>(2.7) | 13.0<br>(3.1) | 14.5<br>(3.1) | 0.424 | **<0.001** | 0.941 | 0.42 | 0.57 |
| Pauses / min | 24.2<br>(2.9) | 26.1<br>(5.1) | 23.9<br>(4.5) | 23.6<br>(3.1) | 0.394 | 0.353 | 0.316 | 0.06 | 0.57 |
| **Gait** |  |  |  |  |  |  |  |  |  |
| Normalized<br>Gait Speed<br>(m/s/m) | 0.65<br>(0.08) | 0.58<br>(0.09) | 0.73<br>(0.12) | 0.69<br>(0.12) | **0.010** | **<0.001** | **0.032** | 0.74 | 1.00 |
| Cadence (steps<br>/ min) | 106.5<br>(8.3) | 102.4<br>(8.5) | 111.7<br>(8.8) | 108.7<br>(8.4) | 0.133 | **<0.001** | 0.099 | 0.60 | 0.73 |

Within the concussion group, longer total pause durations in the ST_S_ condition were strongly associated with greater vestibular-related symptom scores (*ρ* = 0.72) and moderately associated with all other symptom subscores (*ρ* = 0.55-0.66, Table 3). The strength of the associations between symptoms and total pause duration decreased during walking; weak-to-moderate associations were observed for DT total pause duration and DTC_S_ (Table 3) for all symptom scores. Conversely, ST_G_ and DT gait speed was strongly associated with vestibular symptoms (*ρ* = −0.75 and −0.78 respectively, see Table 3 and Figure 2), and moderately associated with all other symptom subscores (*ρ* = −0.50 and −0.63).

**Figure 2.**
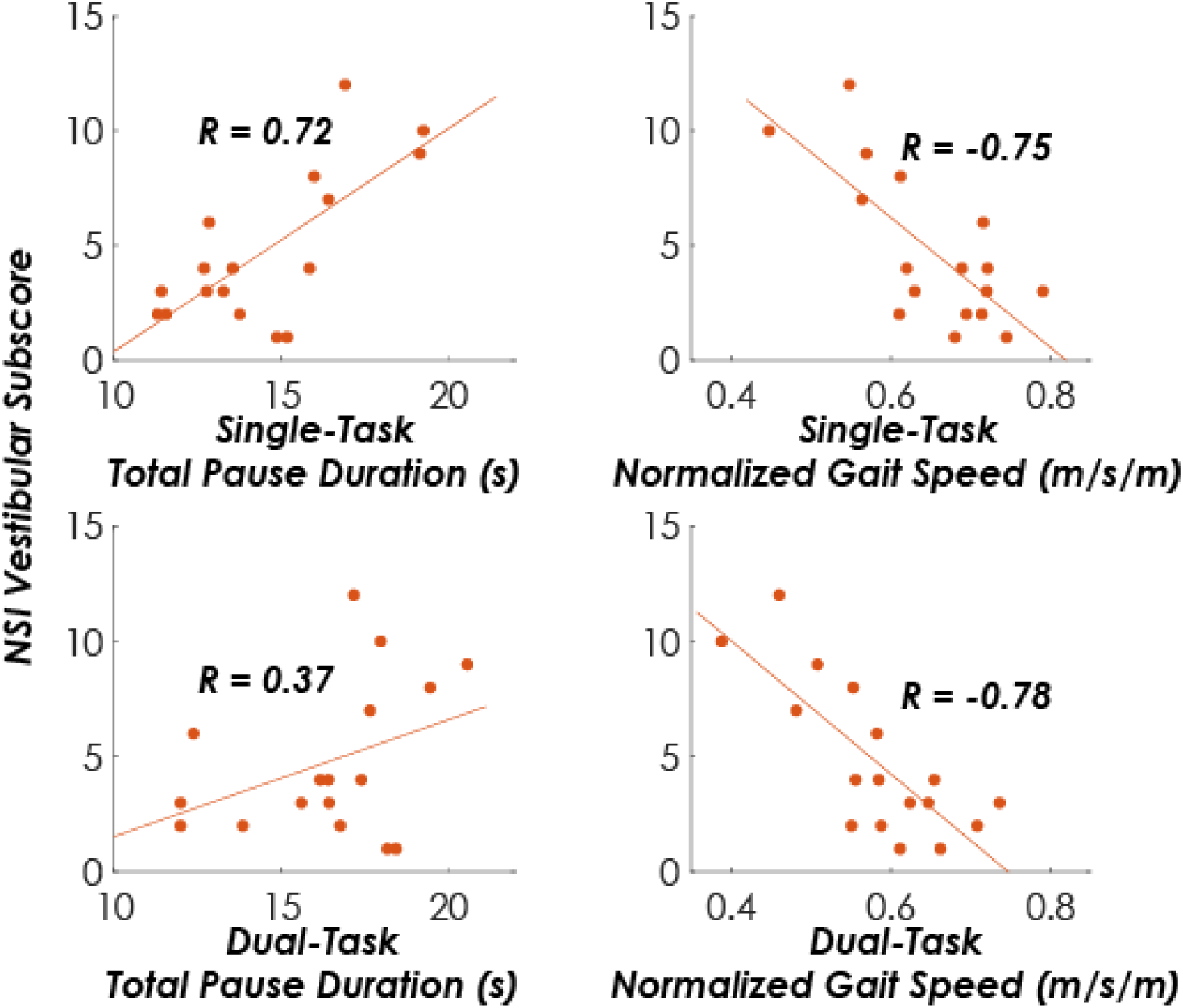
Scatter plots depicting the association between the vestibular subscore on the Neurobehavioral Symptom Inventory with total pause duration and normalized gait speed during single- and dual-task conditions in participants with a concussion.

**Table 3.** Correlation coefficients between self-reported symptoms on the Neurobehavioral Symptom Inventory (NSI) and primary speech (total pause duration) and gait (gait speed) outcomes within the concussion group for single-task (ST), dual-task (DT) and dual-task costs on speech (DTC_S_) and gait (DTC_G_). Bold coefficients indicate strong associations (0.7 – 0.89).

|  |  | NSI-Vestibular | NSI-Cognitive | NSI-Somatosensory | NSI-Affective | NSI-Total |
| --- | --- | --- | --- | --- | --- | --- |
| Total Pause<br>Duration (s) | ST | **0.72** | 0.55 | 0.66 | 0.60 | 0.66 |
|  | DT | 0.37 | 0.26 | 0.41 | 0.30 | 0.37 |
|  | DTC<sub>S</sub> | -0.48 | -0.40 | -0.36 | -0.41 | -0.40 |
| Normalized<br>Gait Speed<br>(m/s/m) | ST | **-0.75** | -0.50 | -0.58 | -0.57 | -0.63 |
|  | DT | **-0.78** | -0.53 | -0.55 | -0.57 | -0.62 |
|  | DTC<sub>G</sub> | -0.45 | -0.34 | -0.21 | -0.29 | -0.30 |

There were no strong associations between any computerized cognitive score and total pause duration or gait speed overall or in any group (Table 4). In the control group, the total duration of speech pauses in ST_S_ and DT conditions had moderate inverse associations with Pattern Comparison Processing Speed in controls (*ρ* = −0.47 and −0.56, respectively), where better cognitive scores were associated with shorter total durations of speech pauses. However, ST and DT total pause duration and Pattern Comparison Processing Speed had only weak inverse associations with one another in people with concussion (*ρ* = −0.32 and −0.28, respectively); instead, ST_G_ speed was moderately associated with Pattern Comparison Processing Speed in those with concussion (*ρ* = 0.43). There were numerous moderate associations between ST_G_ and DT gait speed with all Fluid Cognition cognitive scores, where better cognitive scores were associated with faster ST_G_ and DT gait speed (*ρ* = 0.42 – 0.48, Table 4). Some notable differences in the specific pairs of moderate associations were observed between groups: the concussion group exhibited moderate associations between ST_G_ and DT gait speed with the Dimensional Change Card Sort (*ρ* = 0.45 and 0.48, respectively) while ST_G_ and DT gait speed in controls exhibited very weak associations with Dimensional Change Card Sort (*ρ* = −0.01 and −0.14, respectively).

**Table 4.**
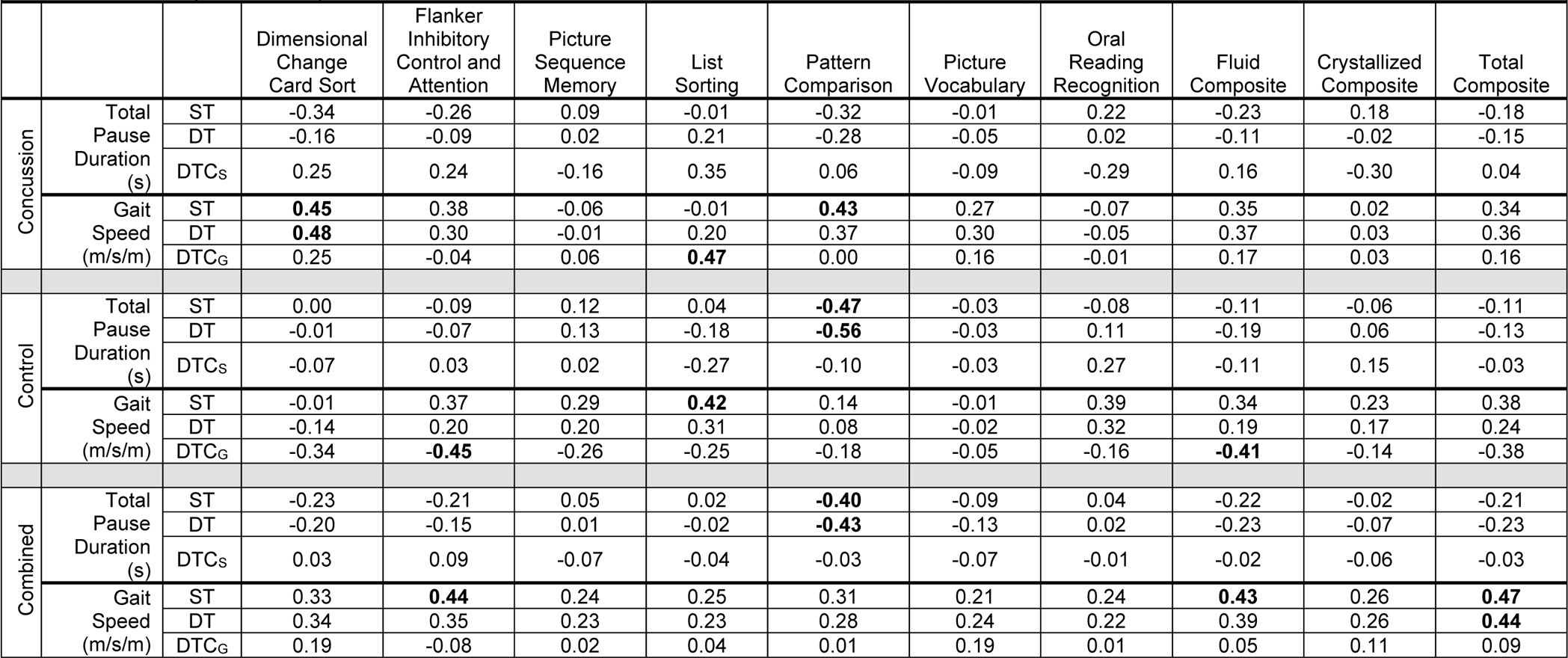
Correlation coefficients between computerized neurocognitive scores on the NIH Toolbox Cognition and primary speech (total pause duration) and gait (gait speed) outcomes within the concussion and control groups, separately, for single-task (ST), dual-task (DT) and dual-task costs on speech (DTCS) and gait (DTCG). All scores from the NIH Toolbox Cognition are fully corrected t-scores. Bold coefficients indicate moderate associations (0.4-0.69)

## DISCUSSION

Our study investigated the effects of concussion on extemporaneous speech production during ST (seated) and DT (walking) paradigms. In agreement with prior work, we observed extemporaneous speech elicited DT costs on walking, indicated by slower gait speeds and slower cadence. Our results did not fully support our hypothesis about group differences; individuals with a concussion did not uniformly exhibit longer pauses during extemporaneous speech and did not demonstrate significantly larger DT costs in speech compared to controls, although moderate effect sizes warrant further study. However, our results indicated that those with a concussion had larger DTC in gait speed than controls. Eliciting greater DTC to gait using an ecologically relevant cognitive DT complements well-established deleterious effects of concussion on DT gait using standardized cognitive tasks ^2,4–7,9,10,13,16^. Further, variable association of DTC with differing symptom domains suggests that the effects will be worst in symptomatic individuals, and more importantly that DT deficits are heterogeneous across people with concussion and may be dependent on symptomology.

While we did not observe any significant group differences in the total duration of pauses or the number of pauses during speech, the effect sizes observed complement prior work suggesting concussion-related symptoms may increase pauses during speech. Prior work in acutely concussed athletes (0-6 days post-concussion) observed increased pause and time fill (i.e., filler word) errors after concussion compared to pre-injury baseline performance ^30^. The difference between these two studies may be attributed to different task demands, symptomology, and time since injury. While prior work used a visual scanning task (describing a picture), the talking task here did not require visual processing. It is possible that speech deficits when describing a picture are compounded by ocular-motor deficits that affect visual scanning. Additionally, participants ranged from 8-14 days post-concussion with varying symptom burdens in the present study. A relatively longer time since injury and the heterogeneous symptom burden at the time of testing revealed a strong relationship between self-reported symptoms, particularly vestibular-related symptoms, and the total duration of pauses during extemporaneous speech. While the mechanisms underlying the association between vestibular symptoms and ST speech, ST gait, and DT gait remain unclear, we speculate that such results originate from attention / rumination on symptoms. Since the strongest associations were with vestibular symptom scores, rather than cognitive scores, we posit that individuals may have been allocating attention to minimize head motion / rotation to avoid exacerbating vestibular symptoms ^40^. Dedicating attention to limiting aversive vestibular stimulation would effectively add a third motor task and another implicit goal to the walking and talking tasks. Further, while we failed to detect statistically significant differences for total speech pause duration, we observed notable between-group effect sizes, especially in DT walking conditions, supporting differences in attention allocation after concussion that warrant further study. Given the associations between vestibular symptoms and our outcomes, future studies should work to understand how concussion affects talking while standing as an intermediary postural challenge to measure cognitive-motor status as a function of vestibular demands (e.g. seated and speaking, standing and speaking, walking and speaking).

While we did not observe a significant group difference in DTC in speech pauses (DTC_S_), we did detect a difference in DTC to walking (DTC_G_), indicated by the group*task interaction, for gait speed. Increased DTC_G_ have been well-documented after concussion, especially in symptomatic individuals within 14 days post-injury ^5^. Previous work used standardized cognitive tasks such as reciting months of the year in reverse, spelling a five-letter word backwards, or serial subtraction, but such tasks lack ecological relevance to daily life. Both cognitive and motor task complexity affect the ability of dual-tasking to differentiate those with concussion from healthy controls ^41,42^, with generally more difficult tasks eliciting larger concussion-related dual-task deficits. Our results extend prior studies and demonstrate that even during ecologically relevant cognitive tasks such as extemporaneous speech, individuals with concussion demonstrate greater motor deficits during a DT compared to healthy controls. However, it remains unclear whether dual-task performance would degrade if extemporaneous speech were paired with more complex motor tasks (e.g., obstacle crossing, turning, etc.). Motor complexity drives task prioritization and the combination of cognitive demand plus added motor challenge could elicit different results ^15,18^. Plummer D’Amato and colleagues (2011) used an obstacle crossing task and revealed that a clock-monitoring task elicited less dual-task costs than spontaneous speech in impaired older adults. Therefore, pairing extemporaneous speech with ecologically relevant gait tasks of increasing motor complexity might further elucidate the impact of concussion-related dual-task deficits on daily life.

Despite the lack of group differences in speech-related outcomes, our results indicate that an extemporaneous speech task elicits a strong dual-task effect overall, in agreement with prior work ^18,23,43^. Both concussion and control groups exhibited greater speech pause durations, slower gait speeds, and slower cadences during the DT compared to the ST condition. Thus, the walking-while-talking task successfully elicited mutual interference on both extemporaneous speech and gait in the majority of participants, regardless of group (see Figure 1). The consistent effect of walking while talking further supports its use as an ecologically relevant cognitive task to assess dual-task gait in young adults.

Results indicating different relationships between cognitive tests and speech pauses or gait speed suggest people with concussion may recruit compensatory cognitive networks during dual-tasking. Here, we observed ST and DT gait speed had moderate, positive relationships with measures of processing speed, cognitive flexibility, and attention, but this relationship was only observed in people with concussion. Associations between gait speeds and cognitive functions mirror those where ST and DT gait speed was associated with measures of associative learning and attention in people with persisting symptoms after concussion, but not in healthy controls ^44^. Our results support prior work suggesting that attention and processing speed is more important for gait in people with concussion compared to controls ^11,41,45^, similar to compensatory recruitment in other mobility-impaired populations ^46,47^. Finally, pauses during an extemporaneous speech task were moderately associated with cognitive processing speed in controls, supporting its validity as a marker of cognitive function.

### Limitations

The primary limitations of this work include the relatively modest sample size and the reliance on silent speech pauses to infer cognitive task performance. Despite the sample size, the consistent chronicity (8-14 days post-injury) and symptomatic nature of the participants is a strength of the study. The reliance on silent speech pauses was grounded in prior work using walking and talking tasks. However, it is possible that examining the syntactic or lexical complexity of the speech may better elucidate differences between groups or tasks compared to only using pauses. Finally, participants were only tested cross-sectionally at a single point in time. Without a baseline from the concussion participants, we cannot determine if their current speech performance was directly caused by the concussion.

## Conclusions

Extemporaneous speech offers an ecologically relevant cognitive DT that elicited larger gait DTC in people after a concussion. While we did not observe significant group differences in speech performance, effect sizes and strong associations between vestibular-related symptoms, total pause duration, and gait speed suggest DTC are variable based on a person’s post-concussion symptomology. Further studies should probe compensatory recruitment of different cognitive processes during DT walking after a concussion, and how such effects may differ based on symptomology. Overall, these results suggest that DT deficits after concussion may have deleterious effects on tasks that are essential to daily life.

## Data Availability

All data produced in the present study are available upon reasonable request to the authors

## ACKNOWLEDGEMENTS

The authors would like to especially thank Cecilia Martindale, Sarah Hill, Dr. Ryan Pelo, Gabrielle Gaudette, Emma Nilsson Read, and Elizabeth Hovenden for their assistance with participant recruitment and data collection on this project.

## REFERENCES

1. Manaseer TS, Gross DP, Dennett L, Schneider K, Whittaker JL. Gait Deviations Associated With Concussion: A Systematic Review. Clin J Sport Med Off J Can Acad Sport Med. 2020;30 Suppl 1:S11–S28. doi:10.1097/JSM.0000000000000537

2. Parker TM, Osternig LR, VAN Donkelaar P, Chou LS. Gait stability following concussion. Med Sci Sports Exerc. 2006;38(6):1032–1040. doi:10.1249/01.mss.0000222828.56982.a4

3. Wood TA, Hsieh KL, An R, Ballard RA, Sosnoff JJ. Balance and Gait Alterations Observed More Than 2 Weeks After Concussion: A Systematic Review and Meta-Analysis. Am J Phys Med Rehabil. 2019;98(7):566–576. doi:10.1097/PHM.0000000000001152

4. Catena RD, van Donkelaar P, Chou LS. Altered balance control following concussion is better detected with an attention test during gait. Gait Posture. 2007;25(3):406–411. doi:10.1016/j.gaitpost.2006.05.006

5. Fino PC, Parrington L, Pitt W, et al. Detecting gait abnormalities after concussion or mild traumatic brain injury: A systematic review of single-task, dual-task, and complex gait. Gait Posture. 2018;62:157–166. doi:10.1016/j.gaitpost.2018.03.021

6. Martini DN, Sabin MJ, DePesa SA, et al. The chronic effects of concussion on gait. Arch Phys Med Rehabil. 2011;92(4):585–589. doi:10.1016/j.apmr.2010.11.029

7. Parker TM, Osternig LR, Lee HJ, Donkelaar P van, Chou LS. The effect of divided attention on gait stability following concussion. Clin Biomech. 2005;20(4):389–395. doi:10.1016/j.clinbiomech.2004.12.004

8. Jain D, Graci V, Beam ME, et al. Neurophysiological and gait outcomes during a dual-task gait assessment in concussed adolescents. Clin Biomech Bristol Avon. 2023;109:106090. doi:10.1016/j.clinbiomech.2023.106090

9. Martini DN, Mancini M, Antonellis P, et al. Prefrontal Cortex Activity During Gait in People With Persistent Symptoms After Concussion. Neurorehabil Neural Repair. 2024;38(5):364–372. doi:10.1177/15459683241240423

10. Büttner F, Howell DR, Ardern CL, et al. Concussed athletes walk slower than non-concussed athletes during cognitive-motor dual-task assessments but not during single-task assessments 2 months after sports concussion: a systematic review and meta-analysis using individual participant data. Br J Sports Med. 2020;54(2):94–101. doi:10.1136/bjsports-2018-100164

11. Howell DR, Kirkwood MW, Provance A, Iverson GL, Meehan WP. Using concurrent gait and cognitive assessments to identify impairments after concussion: a narrative review. Concussion Lond Engl. 2018;3(1):CNC54. doi:10.2217/cnc-2017-0014

12. Bryk KN, Passalugo S, Chou LS, et al. Dual task cost in adults with persistent concussion symptoms. Gait Posture. 2023;101:120–123. doi:10.1016/j.gaitpost.2023.02.008

13. Martini DN, Parrington L, Stuart S, Fino PC, King LA. Gait Performance in People with Symptomatic, Chronic Mild Traumatic Brain Injury. J Neurotrauma. 2021;38(2):218–224. doi:10.1089/neu.2020.6986

14. Yogev-Seligmann G, Hausdorff JM, Giladi N. The role of executive function and attention in gait. Mov Disord Off J Mov Disord Soc. 2008;23(3):329–342; quiz 472. doi:10.1002/mds.21720

15. Yogev-Seligmann G, Hausdorff JM, Giladi N. Do we always prioritize balance when walking? Towards an integrated model of task prioritization. Mov Disord Off J Mov Disord Soc. 2012;27(6):765–770. doi:10.1002/mds.24963

16. Kleiner M, Wong L, Dubé A, Wnuk K, Hunter SW, Graham LJ. Dual-Task Assessment Protocols in Concussion Assessment: A Systematic Literature Review. J Orthop Sports Phys Ther. 2018;48(2):87–103. doi:10.2519/jospt.2018.7432

17. Dromey C, Jarvis E, Sondrup S, Nissen S, Foreman KB, Dibble LE. Bidirectional interference between speech and postural stability in individuals with Parkinson’s disease. Int J Speech Lang Pathol. 2010;12(5):446–454. doi:10.3109/17549507.2010.485649

18. Raffegeau TE, Haddad JM, Huber JE, Rietdyk S. Walking while talking: Young adults flexibly allocate resources between speech and gait. Gait Posture. 2018;64:59–62. doi:10.1016/j.gaitpost.2018.05.029

19. Miyake A, Friedman NP. The Nature and Organization of Individual Differences in Executive Functions: Four General Conclusions. Curr Dir Psychol Sci. 2012;21(1):8–14. doi:10.1177/0963721411429458

20. Lovett MC. A Strategy-Based Interpretation of Stroop. Cogn Sci. 2005;29(3):493–524. doi:10.1207/s15516709cog0000_24

21. Kubose TT, Bock K, Dell GS, Garnsey SM, Kramer AF, Mayhugh J. The Effects of Speech Production and Speech Comprehension on Simulated Driving Performance. Appl Cogn Psychol. 2006;20(1):43–63. doi:10.1002/acp.1164

22. Lee AMC, Cerisano S, Humphreys KR, Watter S. Talking is harder than listening: The time course of dual-task costs during naturalistic conversation. Can J Exp Psychol Rev Can Psychol Exp. 2017;71(2):111–119. doi:10.1037/cep0000114

23. Raffegeau TE, Brinkerhoff SA, Clark M, et al. Walking (and talking) the plank: dual-task performance costs in a virtual balance-threatening environment. Exp Brain Res. 2024;242(5):1237–1250. doi:10.1007/s00221-024-06807-w

24. Mitchell HL, Hoit JD, Watson PJ. Cognitive-linguistic demands and speech breathing. J Speech Hear Res. 1996;39(1):93–104. doi:10.1044/jshr.3901.93

25. Love LR, Starbuck HB, Christensen JM. Pause Frequency in Fluent and Nonfluent Speech. J Acoust Soc Am. 1972;51(1A_Supplement):122. doi:10.1121/1.1981351

26. Krivokapić J, Styler W, Parrell B. Pause postures: The relationship between articulation and cognitive processes during pauses. J Phon. 2020;79:100953. doi:10.1016/j.wocn.2019.100953

27. Lee J, Huber J, Jenkins J, Fredrick J. Language planning and pauses in story retell: Evidence from aging and Parkinson’s disease. J Commun Disord. 2019;79:1–10. doi:10.1016/j.jcomdis.2019.02.004

28. Huber JE, Darling M. Effect of Parkinson’s disease on the production of structured and unstructured speaking tasks: respiratory physiologic and linguistic considerations. J Speech Lang Hear Res JSLHR. 2011;54(1):33–46. doi:10.1044/1092-4388(2010/09-0184)

29. Wang YT, Kent RD, Duffy JR, Thomas JE. Dysarthria associated with traumatic brain injury: speaking rate and emphatic stress. J Commun Disord. 2005;38(3):231–260. doi:10.1016/j.jcomdis.2004.12.001

30. Patel S, Grabowski C, Dayalu V, Testa AJ. Speech error rates after a sports-related concussion. Front Psychol. 2023;14:1135441. doi:10.3389/fpsyg.2023.1135441

31. Wall C, Powell D, Young F, et al. A deep learning-based approach to diagnose mild traumatic brain injury using audio classification. PloS One. 2022;17(9):e0274395. doi:10.1371/journal.pone.0274395

32. Oldham J, Lent B, Peretiatko S, Dec K. Predictors of Speech Language Pathology Referral After Pediatric Concussion Using a Speech Language Checklist. Am J Phys Med Rehabil. 2023;102(10):919–922. doi:10.1097/PHM.0000000000002293

33. Cicerone KD, Kalmar K. Persistent postconcussion syndrome: The structure of subjective complaints after mild traumatic brain injury. J Head Trauma Rehabil. 1995;10(3):1.

34. Hodes RJ, Insel TR, Landis SC. The NIH Toolbox. Neurology. 2013;80(11 Suppl 3):S1. doi:10.1212/WNL.0b013e3182872e90

35. Morris R, Stuart S, McBarron G, Fino PC, Mancini M, Curtze C. Validity of MobilityLab (version 2) for gait assessment in young adults, older adults and Parkinson’s disease. Physiol Meas. 2019;40(9):095003. doi:10.1088/1361-6579/ab4023

36. Bosker HR, Pinget AF, Quené H, Sanders T, de Jong NH. What makes speech sound fluent? The contributions of pauses, speed and repairs. Lang Test. 2013;30(2):159–175. doi:10.1177/0265532212455394

37. Khawaja MA, Ruiz N, Chen F. Think before you talk: an empirical study of relationship between speech pauses and cognitive load. In: Proceedings of the 20th Australasian Conference on Computer-Human Interaction: Designing for Habitus and Habitat. OZCHI ‘08. Association for Computing Machinery; 2008:335–338. doi:10.1145/1517744.1517814

38. Noufi C, Lammert A, Mehta D, et al. Vocal Biomarker Assessment Following Pediatric Traumatic Brain Injury: A Retrospective Cohort Study. In:; 2019:3895–3899. doi:10.21437/Interspeech.2019-1200

39. Hedges LV, Olkin I. Statistical Methods for Meta-Analysis. Academic Press; 1985.

40. Loyd BJ, Dibble LE, Weightman MM, et al. Volitional Head Movement Deficits and Alterations in Gait Speed Following Mild Traumatic Brain Injury. J Head Trauma Rehabil. 2023;38(3):E223–E232. doi:10.1097/HTR.0000000000000831

41. Howell DR, Osternig LR, Koester MC, Chou LS. The effect of cognitive task complexity on gait stability in adolescents following concussion. Exp Brain Res. 2014;232(6):1773–1782. doi:10.1007/s00221-014-3869-1

42. Howell DR, Myer GD, Grooms D, Diekfuss J, Yuan W, Meehan WP. Examining Motor Tasks of Differing Complexity After Concussion in Adolescents. Arch Phys Med Rehabil. 2019;100(4):613–619. doi:10.1016/j.apmr.2018.07.441

43. Plummer-D’Amato P, Altmann LJP, Reilly K. Dual-task effects of spontaneous speech and executive function on gait in aging: exaggerated effects in slow walkers. Gait Posture. 2011;33(2):233–237. doi:10.1016/j.gaitpost.2010.11.011

44. Antonellis P, Weightman MM, Fino PC, et al. Relation Between Cognitive Assessment and Clinical Physical Performance Measures After Mild Traumatic Brain Injury. Arch Phys Med Rehabil. 2024;105(5):868–875. doi:10.1016/j.apmr.2023.10.013

45. Cicerone KD. Attention deficits and dual task demands after mild traumatic brain injury. Brain Inj. 1996;10(2):79–89. doi:10.1080/026990596124566

46. Martin KL, Blizzard L, Wood AG, et al. Cognitive function, gait, and gait variability in older people: a population-based study. J Gerontol A Biol Sci Med Sci. 2013;68(6):726–732. doi:10.1093/gerona/gls224

47. Morris R, Lord S, Bunce J, Burn D, Rochester L. Gait and cognition: Mapping the global and discrete relationships in ageing and neurodegenerative disease. Neurosci Biobehav Rev. 2016;64:326–345. doi:10.1016/j.neubiorev.2016.02.012

